# SARS-COV-2 VIRUS INFECTED PATIENT IDENTIFICATION THROUGH CANINE OLFACTIVE DETECTION ON AXILLARY SWEAT SAMPLES

**DOI:** 10.1101/2021.06.10.21257898

**Authors:** Dominique Grandjean, Capucine Gallet, Clotilde Lecoq-Julien, Riad Sarkis, Quentin Muzzin, Vinciane Roger, Didier Roisse, Nicolas Dirn, Clément Levert, Erwan Breton, Arnaud Galtat, Alexandre Forget, Sébastien Charreaudeau, Fabien Gasmi, Caroline Jean-Baptiste, Sébastien Petitjean, Katia Hamon, Jean-Michel Duquesne, Chantal Coudert, Jean-Pierre Tourtier, Christophe Billy, Jean-Marc Wurtz, Anthony Chauvin, Xavier Eyer, Sabrina Ziani, Laura Prevel, Ilaria Cherubini, Enfel Khelili-Houas, Pierre Hausfater, Philippe Devillier, Loïc Desquilbet

## Abstract

Facing the COVID-19 pandemic, testing individuals in order to promptly isolate positive people is one of the key actions. One approach to rapid testing might be to consider the olfactory capacities of trained detection dogs in order to develop a non-invasive, rapid and cheap mass detection approach, through the Volatile Organic Compounds (VOCs) signature of SARS-CoV-2 infection.

The goal of this study was to determine the individual values of sensitivity and specificity of trained dogs when performing olfactory detection of COVID-19 on axillary sweat samples. Seven dogs were used to sniff a total of 218 samples (62 COVID-19 positive and 156 COVID-19 negative samples), based on a randomised and double-blinded protocol carried out on olfaction cone line-ups. To ensure a wide olfactory range as close as possible to operational conditions, the samples were retrieved from 13 different sites.

Sensitivities varied from 87% to 94% for 6 dogs, and were above 90% for 3 of them. Only one dog, whose sensitivity was 60%, was not selected to subsequently enter the operational stage. Specificities varied from 78% to 92%, with 6 dogs over 85% and 4 dogs over 90%.

Based on the calculated sensitivity and specificity for each dog, positive and negative predictive values (PPV and NPV) were calculated according to several prevalence rates of SARS-CoV-2 scenarii, and were compared to PPV and NPV of an “almost perfect” diagnostic tool. For 6 out of the 7 dogs, and for a prevalence rate of SARS-CoV-2 lower or equal than 40%, the NPV of the dogs were virtually the same as the one of the “almost perfect” tool. Along with other studies on olfactory detection of COVID-19 by dogs these positive and encouraging results suggest that olfactory dogs may play an important part in mass COVID-19 pre-testing situations.

## INTRODUCTION

As the COVID-19 pandemic continues its course in most countries all over the world, the need for fast, reliable and cheap testing methods becomes more obvious as an optimal way of managing the worldwide crisis.

One possible solution lies in the incredible olfactory sense of dogs that can identify the presence of the SARS-CoV-2 in an individual through the signature Volatile Organic Compounds (VOCs) it generates.

The Nosaïs-COVID-19 program, launched by the Alfort School of Veterinary Medicine (France) and the Saint-Joseph University of Beirut (Lebanon) at the start of March 2020, aims at developing the use of dogs’ olfactory abilities in detecting the SARS-CoV-2 carriage of asymptomatic or paucisymptomatic individuals through smelling axillary sweat sample, under controlled conditions. An initial proof-of-concept study led by our team provided some evidence that this approach had conclusively shown the existence of a specific sweat scent in COVID-19 positive individuals (via the RT-PCR SARS-CoV-2-IP24 technique of the Pasteur Institute, recommended by the World Health Organisation [1, 2] which seems to be identified by detection dogs [3]. Articles from other teams followed our proof-of-concept study, showing similar results with saliva [4], respiratory secretions [5, 6], and urine samples [7].

Now that the preliminary work had been done, we were tasked with setting up the validation stage of the concept with randomised tests to produce accurate individual values of sensitivity and specificity for the canine olfactory test, which is the subject of this study.

For some years now, it has been written that VOCs have the potential to become a revolutionary and non-invasive approach to medical diagnostic in humans. This led to wondering whether the viral infection could activate cellular production of specific molecules, with varying degrees of detection difficulty, specific to the studied virus, or the direct release of specific catabolites by the virus. Several studies focused on the VOCs produced by the infection process, but all were linked to the inflammatory consequences of the oxidative stress induced, and no further research was done towards the characteristics related to the relevant pathogenic agent [8-13]. In 2012, Aksenov et al. studied the VOCs produced by cultures of B-lymphocytes infected by three influenza viruses: avian H9N2, avian H6N2 and human H1N1 [14, 15]. The families of collected VOCs turned out to be unique and specific to each viral subtype. The authors concluded that the minor alterations induced by the virus on the cell’s genome led to a specific change in the production of VOCs in the cellular metabolism. Recently, Abd El Qader et al. showed the specificity of bacterial or viral species in the VOCs produced by infected cell cultures [16]. A review of 30 articles on the subject found that most bacteria produce species specific VOCs can help as a diagnostic basis for infected patients [17]. Schivo et al. also identified a specific volatilome on airway cells infected by a rhinovirus [18].

The hypothesis supported by these studies is the generation of VOCs specific to the SARS-CoV-2 virus in the infected cell which determine the presence of an active virus in the human organism, when they are detectable.

Angle et al. noted in a recent review in 2016 that these VOCs’ exit from the body and that, at that time, no study had been done on the subject [19]. They stated that for the use of canine olfactory detection, there were two possible options: targeted “scenting” (from a natural emunctory) and “plume scenting”, which is the global olfactory analysis concept used by dogs, through the thermal and aerodynamic flux that carry scent from the human body. This last option is not a new concept, as it is used by search and rescue dogs to find people in disaster and area searches. All over the world, we continue to develop the use of dogs’ noses in the search for explosives, drugs, bank notes, as well as animal trafficking [20].

For centuries, even our own sense of smell was put to use by doctors in their everyday work, whether it was to recognise gangrene on a battlefield or identify the ketoacidosis stage of diabetes. From there, canine olfactory detection has been the subject of a number of studies, many of which have shown a similar, sometimes even better, accuracy but mostly earlier detection than standard analytic methods.

In 1989, Williams and Pembroke introduced the idea that a dog might be able to detect a malignant tumour via the specific scent it produces [21]. Willis et al. published the first clinical inquiry related to cancer, specifically bladder cancer [22], following a proof-of-concept study published in 2004 [23]. Several studies followed with positive results on olfactory detection of VOCs by dogs for early testing of colorectal cancer [24, 25], and encouraging results for lung cancer [26-29], melanoma [30, 31], prostate cancer [32-34] and liver cancer [35]. Pirone and Albertini recently published a systematic review of the existing literature on the subject [36]. It is also important to note that dogs’ olfactory senses are commonly used for the prevention and support of crises in diabetic [37-40] and epileptic [41] patients.

The use of canine olfactory detection is expanding greatly in the field of infectious diseases and parasitic infestation. This allowed Wallner and Ellis to train dogs to find Gipsy moth eggs with a sensitivity of 95% [42] and Richards et al. proved a dog’s ability to identify sheep suffering from nematode infestation [43]. More recently, Guest et al. showed that some dogs identified people with malaria [44]. Some bacterial diseases, like *Clostridium difficile* infections can also be identified by dogs [45, 46]. Another study successfully showed that it was possible to distinguish, by canine olfactory detection, cell cultures infected by different viruses (bovine *mucosae disease*, bovine herpes 1 or bovine para influenza 3) [19].

Thanks to constant scientific progress, we are now aware of the potential offered by canine olfactory detection in the field of human disease screening. This is what encouraged the Nosaïs team (canine olfactory detection group) from the Alfort School of Veterinary Medicine to develop a scientific program about dogs as a potential non-invasive, fast and cheap tool in operations deployed to test and fight COVID-19.

## MATERIALS AND METHODS

Our study was carried out in strict accordance with the recommendations published in the Guide for the care and use of animals edited by French laws (articles R214-87 to R214-137 of the rural code), updates by decree 2013-118 and five decrees edited on February 1^st^ 2013. The protocol was approved by the committee on the ethics of animal experiments of the Ecole Nationale Vétérinaire d’Alfort (Alfort School of Veterinary Medicine) and by the Protection of Persons Committee (CPP) of the Ile-de-France on march 30^th^ 2020. It also follows the rules of the French Public Health Code (article L1121-1/2). This research is part of the study “COVIDEF” promoted by Assistance Publique - Hopitaux de Paris (Parisian hospitals, AP-HP) led by Professor Hausfater, as “COVIDOG” (Cohort of patients infected by the virus SARS-CoV-2 or suspected to be infected), for the samples coming from AP-HP, and of the study « VOC-COVID-Diag » promoted by Foch hospital (Suresnes) led by Professor Devillier. The latter study has been approved by the CPP Nord Ouest IV on October 8^th^ 2020.

### Recrutement of patients

COVID-19 positive and negative patients were recruited from 13 centres, COVID-19 screening centres, or fire department centres around Paris, France: Hôpital d’Instruction des Armées Bégin (Saint-Mandé), Hôpital Henri Mondor (Créteil), Hôpital de La Pitié Salpêtrière (Paris), Hôpital Lariboisière (Paris), Hôpital Foch (Suresnes), Centre Hospitalier de Rambouillet (Rambouillet), Centre Hospitalier François Quesnay (Mantes-la-Jolie), Centre de Secours de Marly-le-Roi (Marly-le-Roi), Centre Hospitalier Intercommunal Meulan Les Mureaux (Meulan-en-Yvelines), Centre Hospitalier de Poissy (Poissy), Centre hospitalier Saint-Morand (Altkirch), Centre de Secours de Houilles (Houilles), Centre Hospitalier de Saint-Germain en Laye (Saint-Germain En Laye), and Centre de Secours de Chatou (Chatou).

COVID-19 positive and negative axillary sweat samples used for the training and the testing sessions were collected by doctors and nurses in these hospitals and centres, who were trained not to contaminate the samples with their own odours. The reasons for choosing sweat, the sampling site, the method, and the biological safety measures have been explained in a previous publication [3].

Patients presenting to one of the participating hospitals or centres with COVID-19 clinical symptoms (such as fever, cough, throat pain, fatigue or body pain), and having a positive Reverse Transcription Polymerase Chain Reaction (RT-PCR) or PCR test for SARS-CoV-2 were included as COVID-19 positive individuals. To avoid potential interferences with long-term medical treatments in the sweat VOCs, COVID-19 positive individuals were not included in the study if they had received a medical treatment for more than 36 hours prior to the test sampling.

The avoid potential confounding due to “hospital background odour” [36, 47], COVID-19 negative individuals were recruited from the same hospital as COVID-19 positive individuals (i.e., COVID-19 positive and negative individuals were matched on hospital). Inclusion criteria included: being hospitalised the same day of the COVID-19 positive individual for reasons other than COVID-19 symptoms with a negative COVID-19 PCR test result, or being physicians or nurses working in the same hospital who tested negative for COVID-19 within the previous 48 hours. All individuals (COVID-19 positives and negatives) meeting these inclusion criteria were asked if they were willing to participate in the study and signed an individual informed consent form approved by the national ethics committee.

### Samples

As previously presented [3], our assumption was based on the potential excretion of specific catabolites in the sweat, induced by SARS-CoV-2 cellular actions or replications in the organism’s cells, through the apocrine sweat glands, generating VOCs that the dog can detect. Most recently, VOCs have received considerable interest to obtain insights into physiological and physio-pathological processes, and to exploit the knowledge of their absence/presence or changes in their concentration profiles or VOC composition in various body matrices for disease detection and therapeutic monitoring [48-50]. A major advantage of VOCs in this regard is the fact that they are readily and noninvasively obtainable and could be sampled as often as desired with no discomfort [51].

VOCs are volatile at an ambient temperature, and may be detectable by dogs in breath, urine, tears, saliva, faeces and sweat. Most studies on volatile biomarkers have been conducted on breath samples, but their use seems too risky regarding SARS-CoV-2 infection. VOCs emanating from the skin contribute to an individual’s body odour, and may convey important information about metabolic processes [52, 53]. They are produced by eccrine, apocrine and sebaceous gland secretions, and are the major source of underarm odorants, playing a role in chemical signaling [54]. Several studies have been performed to better characterise VOCs produced by axillary apocrine glands [55-58]. Sweat from the palms of the hands, soles of the feet and the forehead mainly comes from eccrine glands and sebum. Most of these sweat compounds are organic acids ranging in carbon size from C2 to C20, the most abundant being saturated, monounsaturated and di-unsaturated C16 and C18, which are not volatile at body temperature [59]. Most recent studies confirmed that human sweat compounds differ according to the anatomic site [60], and that the secretory capacity of eccrine sweat glands appears larger on the trunk (including under the arms) compared to other parts of the body [61].

For the reasons mentioned above, we decided in our previous study to collect axillary sweat [3], which is also a key odour for the search and rescue [62, 63], and the axillary region is easily accessible. Furthermore, this site is unlikely to be contaminated by saliva of a COVID-19 positive patient. However, since it seems, albeit not clearly demonstrated, that sweat is a SARS-CoV-2 transmission route via skin-to-object to-mucosa contact [64], careful manipulation of axillary sweat samples was required. The sampling material used were 2 x 2-inch sterile gauze swabs used by the hospitals. Swabs remained in contact with the armpit skin for ten minutes and the average amount of sweat obtained was around 75mg per swab.

Sweat samples were collected by doctors, interns and nurses by placing two sterile compresses under the armpits of each patient. Sampling protocol began by the explanation of the research project to the participating patients and was followed by the signature of an informed consent form by the patient.

Samplers were trained to collect samples through a training video: they had to wear two pairs of disposable gloves and protective equipment against SARS-CoV-2 when they collected samples from COVID-19 positive patients. For all COVID-19 positive and negative patients, samplers wore disposable gloves as not to pollute samples with their own odour. Each hospital had its own gloves, identical for the sampling of COVID-19 positive and negative patients.

Samples were placed and stored in medical anti-UV sterile glass containers, disinfected by the sampler’s helper, anonymously coded, then placed into a second plastic envelope. Individual anonymous data were registered on a form for each coded sample.

Medical staff recorded demographic and medical data about the recruited patients. The demographic data included: age and sex. The medical data included clinical signs (dyspnoea, fatigue, fever, dry cough, muscular pain, headache, loss of smell, loss of taste, diarrhoea, nasal discharge, colic and migraine), previous or current history of diseases (hypertension, diabetes, overweight, disease and arthrosis) and drugs used at the time of sampling (painkillers, anti-coagulant, anti-hypertension, anti-inflammatory, antibiotics, anti-acid, anti-diabetes, anti-cholesterol, bronchodilator, anti-depressing, thyroid hormone, anxiolytics, and anti-spasmodic).

All samples were transferred from the sampling site to the testing site in separated coolers which were cleaned and disinfected with a 10% aqueous acetone solution after each use. All samples were stored in a refrigerator at a temperature of +4°C (but positives and negatives were not mixed), and were never manipulated without disposable surgical gloves to prevent contamination.

During training and testing processes, the quality of the samples was essential, which justified this detailed description.

### Canine resources and initial training

The dogs trained for this test study were 7 operational search and rescue dogs (Table 1) from French fire departments (Service Départemental d’Incendie et de Secours 78 et 60). These dogs were previously trained to perform area and disaster searches based on human scent.

**Table 1.** Characteristics of the 7 COVID-19 sniffing dogs.

| Name | Gender | Breed | Age (years) | Organisation | Speciality |
| --- | --- | --- | --- | --- | --- |
| Leyko | M | Malinois | 5 | SDIS78* | SAR**** |
| Jinko | M | Groenendael | 6 | SDIS78 | SAR |
| Ortie | F | Malinois | 2.5 | SDIS78 | SAR |
| Oska | F | Malinois | 2.5 | SDIS78 | SAR |
| Oxmo | M | Malinois | 2.5 | SDIS78 | SAR |
| Ouija | M | Dutch Shepherd | 2 | ENVA** | None |
| Joye | F | Malinois | 6 | SDIS60*** | SAR |

\* Service Départemental d'Incendie et de Secours 78 (Fire and Rescue Service 78)
\*\* Ecole Nationale Vétérinaire d'Alfort (Alfort School of Veterinary Medicine)
\*\*\* Service Départemental d'Incendie et de Secours 60 (Fire and Rescue Service 60)
\*\*\*\* Search and Rescue Dog

They were trained to detect SARS-CoV-2 during 8 weeks with line-ups of olfaction cones (figure 1), positive marking by sitting in front of the cone (figure 2) and based on positive reinforcement.

**Figure 1.**
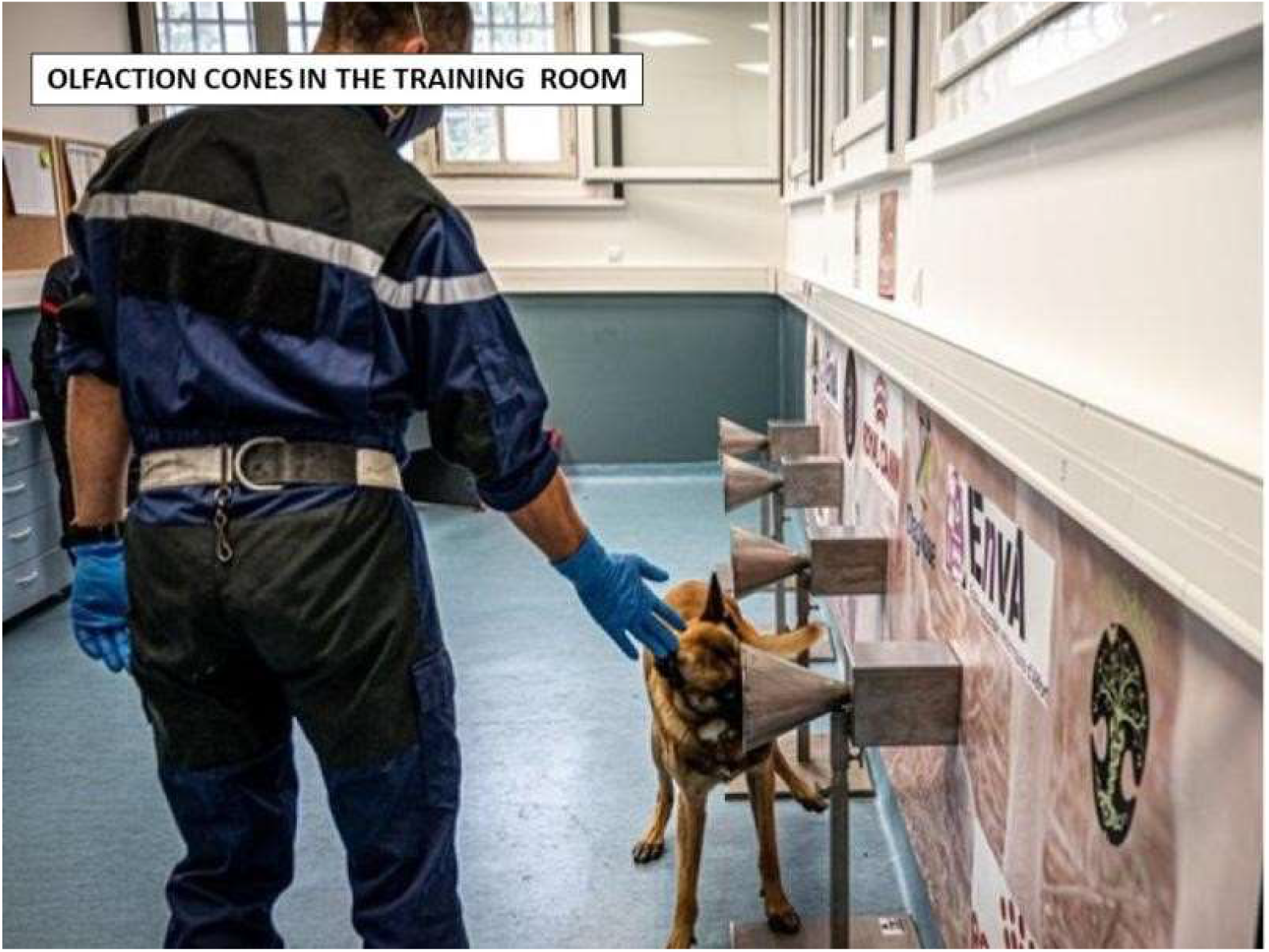
Line-up of olfaction cones in the training room.

**Figure 2.**
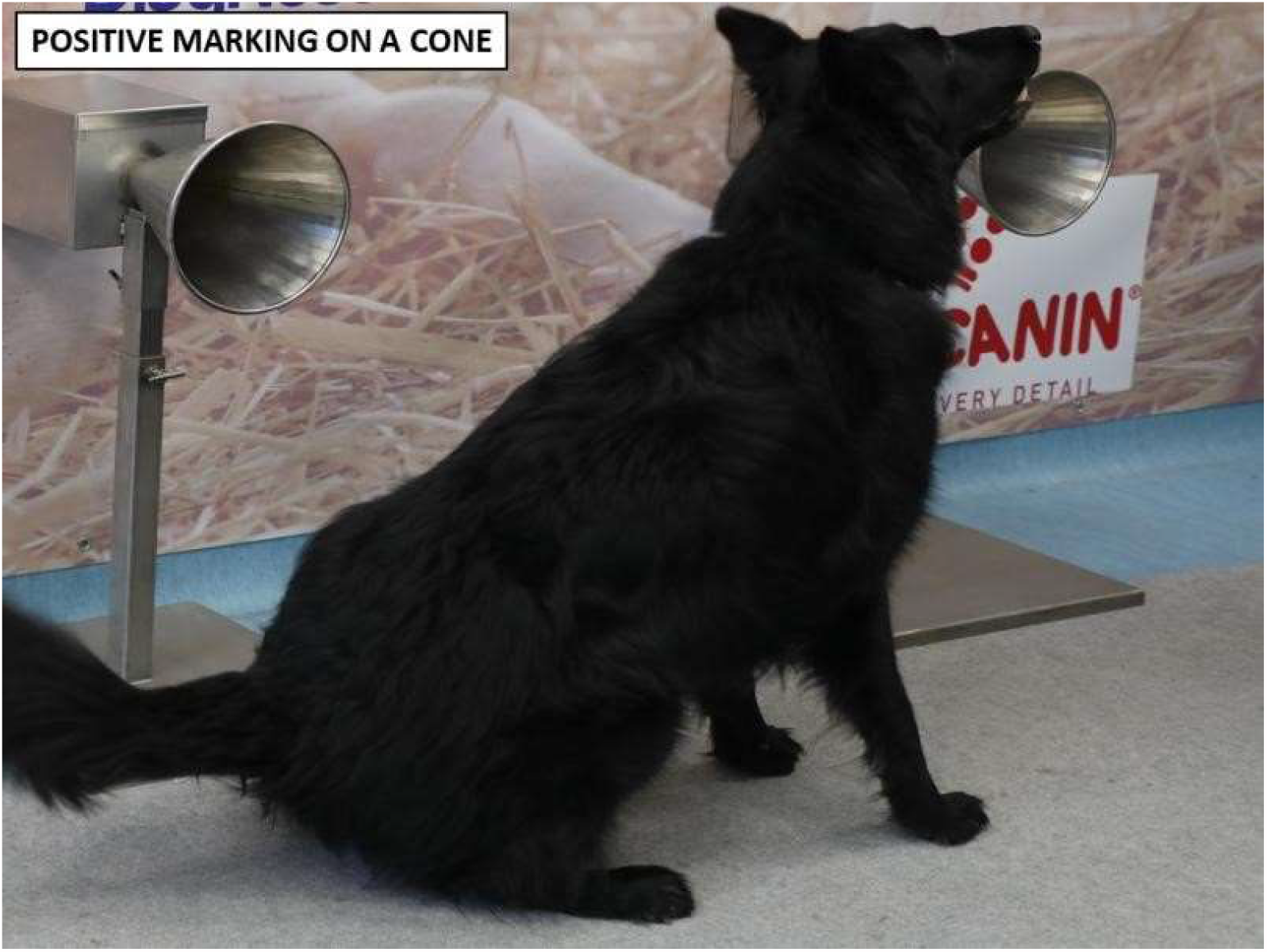
Positive marking of a dog, sitting in front of a cone containing a positive sample.

The training started on September 14^th^ 2020 for the first trained dog (Joye) and ended on November 6^th^ 2020 for the last trained dog (Ortie). The training process followed a five-step procedure: learning line-up work with olfaction cones (step #1), imprinting (memorization) of COVID-19 samples odour (positive samples and empty cones in the line-up; step #2), introduction of mocks (positives sample and virgin swabs in the line-up; step #3), introduction of negatives with no more mocks in the line-up (step #4), and removing all positive samples (only negative samples in the line-up; step #5). Based on the dog’s behaviour, the handlers judged when each dog was ready for the validation testing session. All 7 dogs were deemed trained and ready for the testing session after the 8-week training.

The welfare of the dogs was fully respected, with toy rewarding, and no physical or mental fatigue induced. A total of 106 positives and 242 negative different samples were used during training.

### Biological safety of dogs and humans

There have been very few reports on dogs being passive carriers or very mild symptoms involving SARS-CoV-2. Idexx laboratories, in the USA, tested more than 3500 dogs, cats and horses from places where community transmission of SARS-CoV-2 was occurring in humans and found no positive animals [65]. A recent study performed in Alfort School of Veterinary Medicine (France) provided some evidence of absence of SARS-CoV-2 infection in dogs in close contact with a cluster of COVID-19 patients [66]. Finally, both the CDC (Centers for Disease Control and Prevention, USA), the OIE (International Animal Health Organization) and the ANSES (French Agency for food, environmental and occupational health and safety) attest that there is no evidence that pet animals, and especially dogs, play any significant role in SARS-CoV-2 transmission or spread [66-68].

It is assumed that, like SARS-CoV-1, SARS-CoV-2 does not survive longer than a few hours on cotton [69] which is why, for safety reasons, the samples (stored at +4°C) were not used for training or testing sessions within 24 hours of collection. A more recent study concludes that absorbent materials like cotton are safer than unabsorbant materials for protection from SARS-CoV-2 infection [70]. Finally, the dogs were never in contact with any samples thanks to the design of the olfaction cones (figure 3).

**Figure 3.**
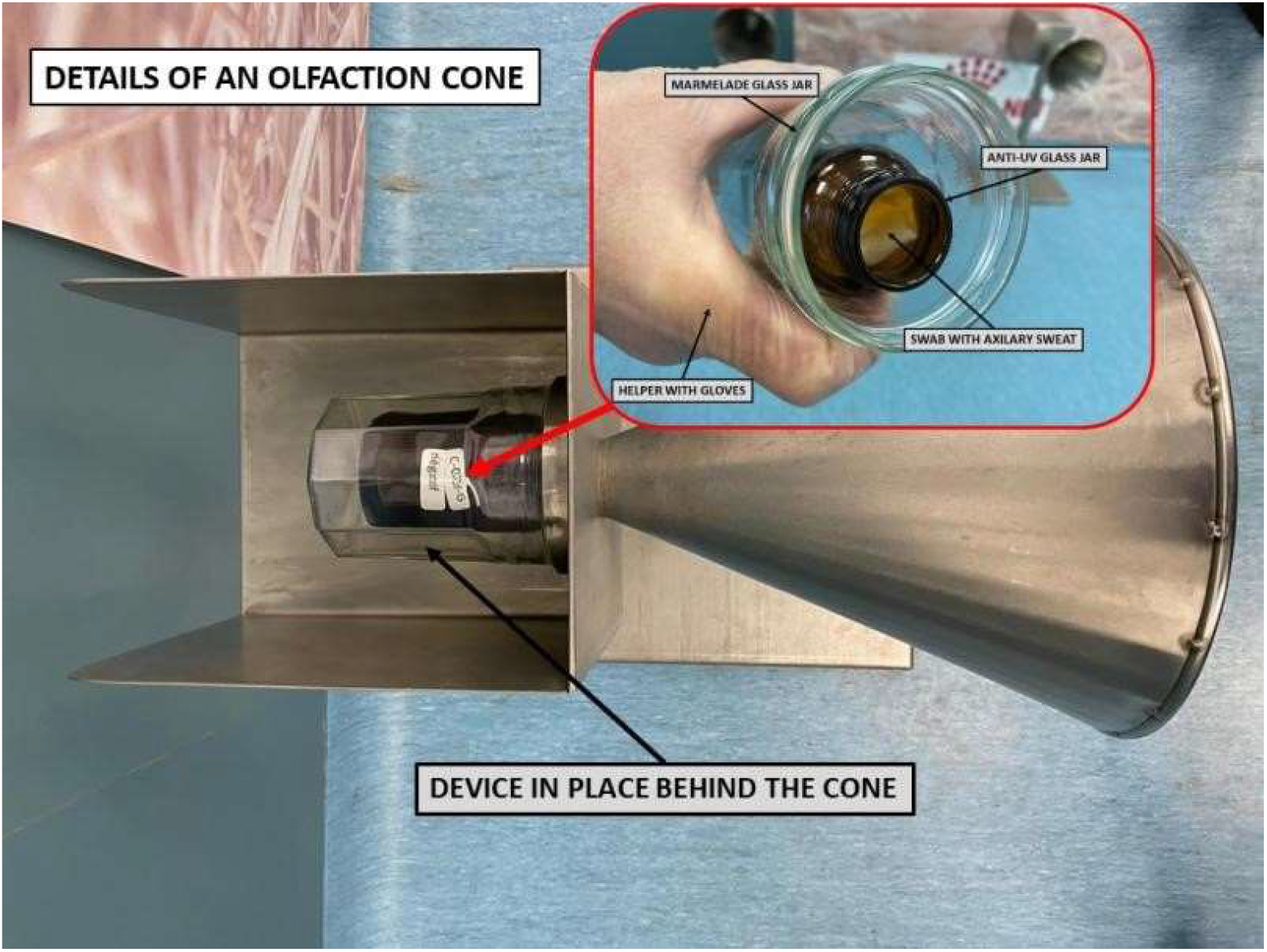
Details of olfaction cone, with doubled protected sample and no possibility of direct contact with the dog.

### Testing protocol

The testing sessions took place in a dedicated room (figure 4) in the rescue center of Poissy (Fire Department 78). A line-up made of 5 to 8 olfaction cones was placed in the room. During trials, all the cones contained COVID-19 negative and/or positive samples, and both the number of positive and negative samples and their position in the line-up were randomised, as recommended by Johnen [71], using a dedicated website (http://www.randomization.com), and double-blinded to the handler and his dog and to the data-recorder. The samples were placed in their position by a dedicated person, while the room was empty, and this person immediately left the room without any contact with other people. After the placement of each sample in the line-up, the data recorder entered in the room and sat behind a one-way glass in order not to be seen by the dog or his handler (figure 5). When a dog entered the room with his/her handler, it had to sniff each cone one by one, as it was trained to do. Samples considered positive by the dog were marked by sitting, barking or scratching in front of the concerned cone; the dog was then rewarded (whether the marking was correct or not) by the handler, who announced the marking to the data-recorder. The handler then asked the dog to resume the task for the remaining cones in the line-up (i.e., sequential line-up). Only when all the dogs had completed a line-up were dog handlers informed of the COVID-19 positive sample(s)’ location in the line-up.

**Figure 4.**
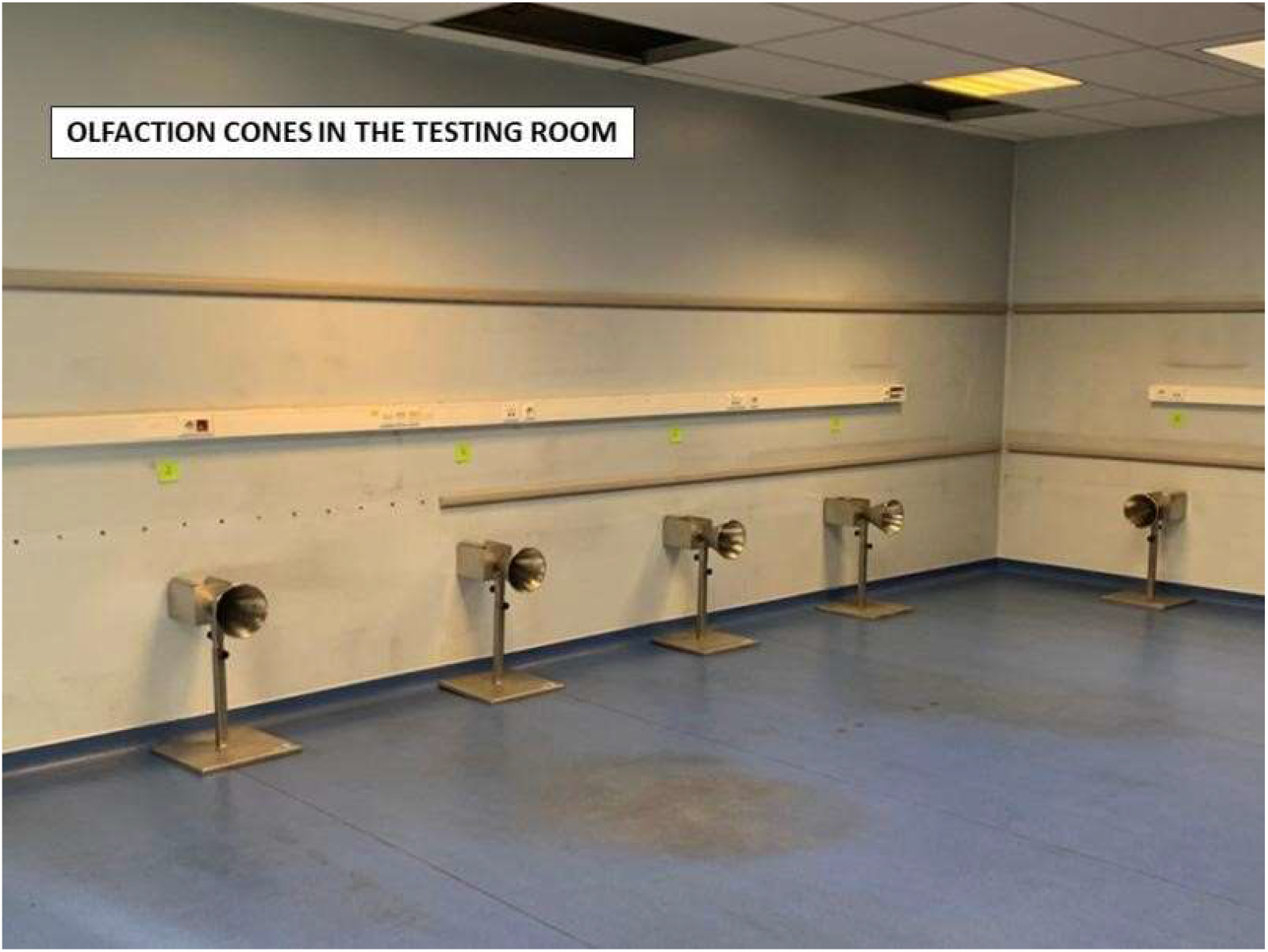
Testing room with its olfaction cones.

**Figure 5.**
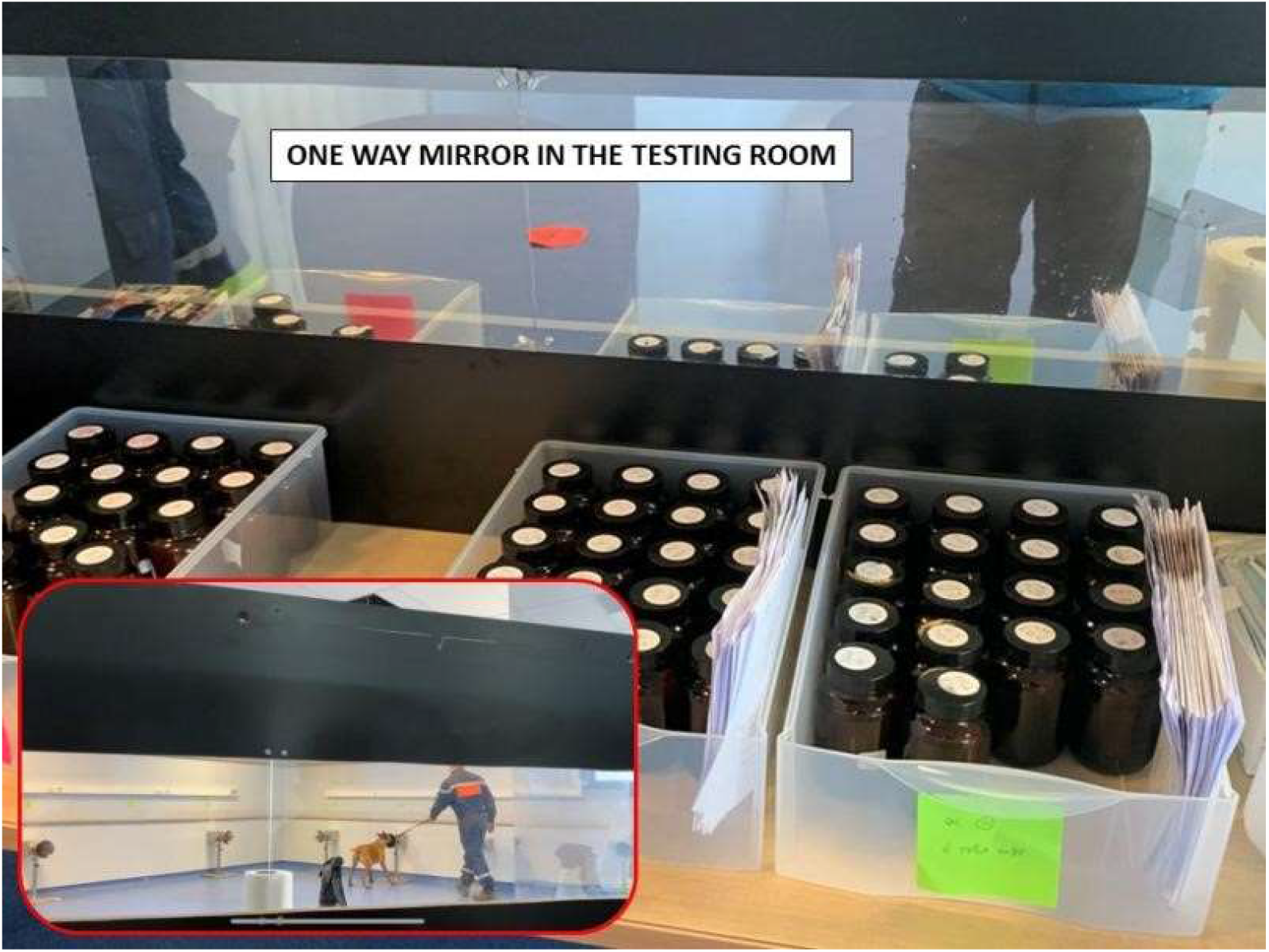
One way glass hiding the recorder in the testing room.

A total of 30 line-ups were performed by all the 7 dogs (except one dog who performed 28 line-ups because of an emergency mission). Once a dog had sniffed all the cones of the line-up, the dog-handler dyad left the room, and the cones were rapidly cleaned by the data recorder with clear water, in order to remove traces left by the previous dog. The next dog then entered the room and sniffed the cones of the same line-up.

Once the 7 dogs had performed the line-up, the cones were cleaned with 3% concentrated acetone solution, new randomised samples were placed in the cones ten minutes after cleaning (neither the handlers, dogs or data recorder were in the room during sample placements, as previously mentioned) and a new trial cycle could start. None of the samples used for the testing sessions had ever been used during the training sessions, and no dog ever sniffed a sample more than once.

For each new sample placements, the person in charge had to wear new disposable gloves (of the same brand during the entire period of testing sessions) and a mask in order not to contaminate the olfactive environment. Out of the 30 line-ups, most consisted of 8 cones (73%; Table 2). The described randomisation of the distribution of the samples in a line-up was designed to ensure that each line-up contained at least one positive and one negative COVID-19 sample. Table 2 describes the number of COVID-19 positive and negative samples among the 30 test line-ups.

**Table 2.** Characteristics of the 30 line-ups used for testing sessions.

|  | Overall (n=30) |
| --- | --- |
| **Number of cones, n (%)** |  |
| 5 cones | 6 (20) |
| 6 cones | 2 (7) |
| 8 cones | 22 (73) |
| **Number of COVID-19 negative samples in the line-up, n (%)** |  |
| 3 | 3 (10) |
| 4 | 6 (20) |
| 5 | 6 (20) |
| 6 | 12 (40) |
| 7 | 3 (10) |
| **Number of COVID-19 positive samples in the line-up, n (%)** |  |
| 1 | 6 (20) |
| 2 | 17 (57) |
| 3 | 6 (20) |
| 4 | 1 (3) |

### Statistical analysis

Sensitivities (Se) were calculated for each dog by dividing the number of COVID-19 positive samples correctly marked by the dog by the total number of COVID-19 positive samples sniffed by the dog during the testing sessions. Specificities (Sp) were calculated for each dog by dividing the number of COVID-19 negative samples which were not marked by the dog by the total number of COVID-19 negative samples sniffed by the dog during the testing sessions.

In order to remove potential confounding bias (i.e. high Se and Sp due to specific odour of COVID-19 positive and negative individuals not related to the presence or absence of SARS-Cov-2 [36, 72]), Se and Sp were also calculated separately in males, females, individuals younger and older than 50 years old, patients recruited from Foch and Rambouillet hospitals (hospitals from which the recruitment of COVID-19 positive patients was the highest), among samples of individuals who did not present the most frequently reported clinical signs (dyspnoea, fatigue, fever, dry cough, and muscular pain), among those who did not have the most frequently reported past or current diseases (hypertension and diabetes), and among those who were not treated by the most frequently reported drugs used (painkillers, anti-coagulant, anti-hypertension, anti-inflammatory, and antibiotics). The 95% confidence interval (CI) was calculated using Jeffreys’ method [73]. Positive and negative predicted values (PPV and NPV, respectively) were calculated for each dog based on their observed Se and Sp and according to three scenarii regarding the risks that an individual is likely to be infected by SARS-CoV-2 [74, 75], based on clinical symptoms, prevalence rate of SARS-CoV-2 in the housing or working area, ranging from 10% to 50% of risk. This range of possible values of probabilities of being SARS-CoV-2-infected is the expected one in settings where COVID-19 sniffing dogs are likely to work (such as in airports or at the entrance of mass events). Positive and negative predicted values are also presented for an almost perfect reference diagnostic tool (with 95% for both Se and Sp) in order to compare the performances of the dogs with this almost perfect diagnostic tool.

Binary and qualitative variables were presented as numbers and proportions, and the quantitative variable (age) was presented as medians and interquartile ranges. Statistical analyses were performed using SAS® University Edition (SAS Institute Inc., Cary, NC, USA).

## RESULTS

A total of 62 COVID-19 positive patients and 156 COVID-19 negative patients were recruited for this study, producing 218 sweat samples used in testing sessions. The proportion of females was slightly higher among COVID-19 negative patients (50%) compared with COVID-19 positive patients (44%; Table 3). The distribution of age was similar between the two groups (medians of 50 and 56 years old, respectively for the COVID-19 negative and positive patients). Patients were recruited mainly from Foch hospital (17%), from Rambouillet hospital (16%), and from the Centre de Secours principal de Houilles (Main Rescue Centre of Houilles) (15%; Table 3). Data about hospitals were missing for 16% of the recruited patients, mainly because of the dispersion of the recruitment in a large number of hospitals where samplers did not fill in the requested documents correctly. The most frequently reported clinical signs were dyspnoea (19%), fatigue (16%), fever (14%), dry cough (14%) and muscular pain (12%), regrouped on Table 3. The most frequently reported past or current diseases were hypertension (26%) and diabetes (13%; Table 3). Each of these clinical signs and past or current diseases were more frequent among COVID-19 positive patients than with COVID-19 negative patients. Other reported clinical signs included headache (7%), loss of smell (6%), loss of taste (6%), diarrhoea (4%), nasal discharge (1%), colic (1%) and migraine (1%). Other reported past or current diseases included overweight (7%), hypercholesterolemia (6%), cancer (6%), chronic kidney disease (5%), asthma (4%), hypothyroidism (4%), heart failure (3%), Parkinson disease (2%), and arthrosis (1%). The most frequently reported drugs used were painkillers (19%), anti-coagulant (15%), anti-hypertension (13%), anti-inflammatory (11%), and antibiotics (10%) (Table 3). Other reported drugs used included anti-acid (5%), anti-diabetes (5%), anti-cholesterol (4%), bronchodilator (2%), anti-depressing (2%), thyroid hormone (2%), anxiolytics (1%) and anti-spasmodic (1%).

**Table 3.**
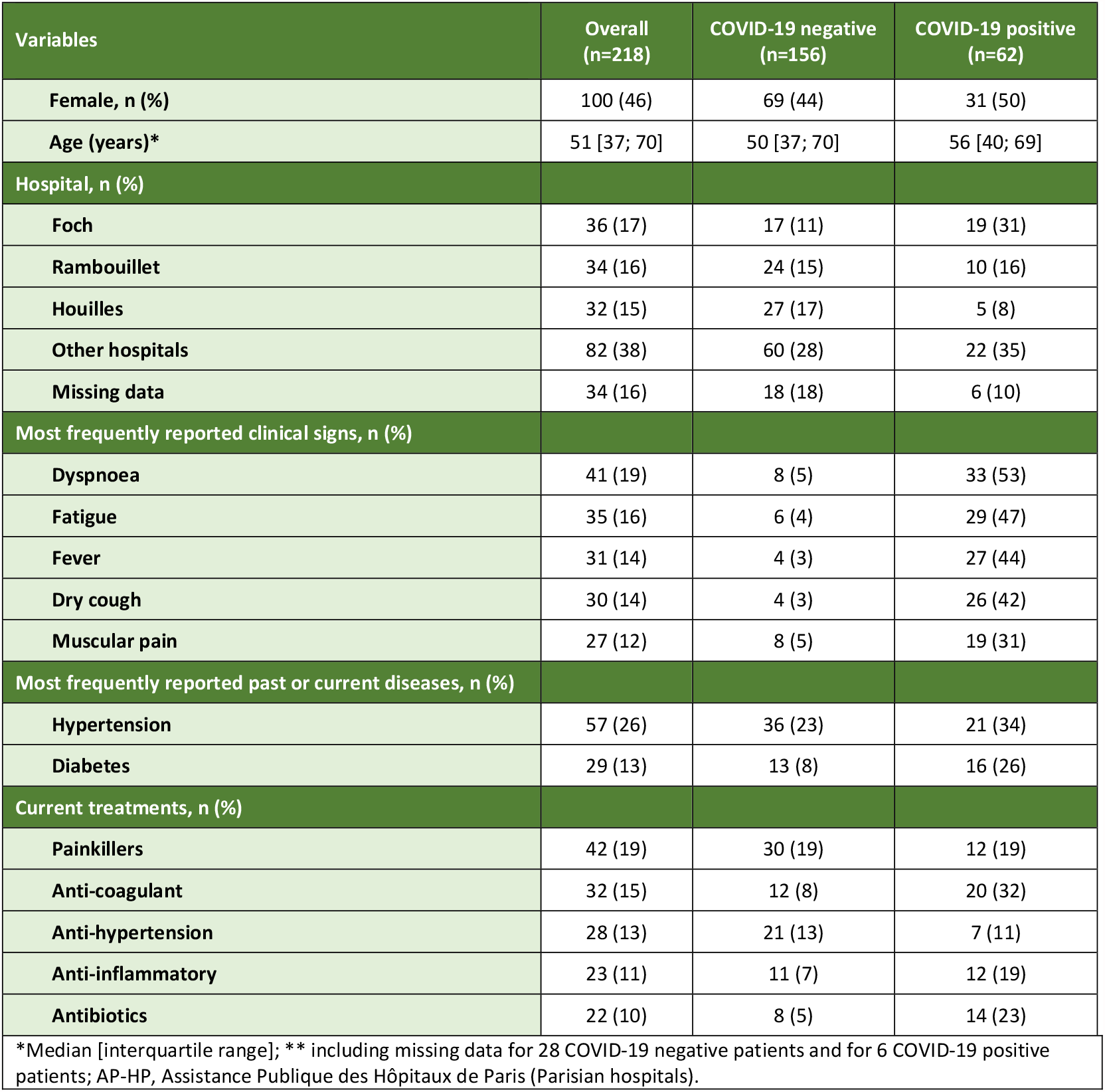
Baseline characteristics of the 218 COVID-19 positive and negative patients.

The overall Se calculated from samples of patients recruited for this study were all equal or higher than 87% except for one dog (Ortie), where Se was 60%. The overall Sp were all equal or higher than 85% except for one dog (Joye), where Sp was 78% (Table 4). Sensitivities remained virtually the same after stratification for sex and age (≤ versus > 50 years old), for patients recruited from hospitals (Foch or Rambouillet hospitals), for those who were free of the most frequently reported health conditions, and for those who did not use the most frequently reported drugs (Tables 5 and 6). After restricting by hospitals, sensitivities were close to the overall numbers, but because of the small number of COVID-19 positive patients in each of the two hospitals, sensitivities may have lacked some accuracy. The sensitivities calculated after restricting patients to those free of diabetes were slightly lower than the overall sensitivities, although all but one remained equal or higher than 85% (57% for Ortie). Specificities remained virtually the same after stratification for sex and age (≤ versus > 50 years old), for patients who were free of the most frequently reported health conditions, and for those who did not use the most frequently reported drugs (Tables 5 and 6). The specificities calculated after restricting to patients recruited from Foch hospital were slightly lower than the overall specificities, which was however not the case after restricting to patients recruited from Rambouillet hospital.

**Table 4.**
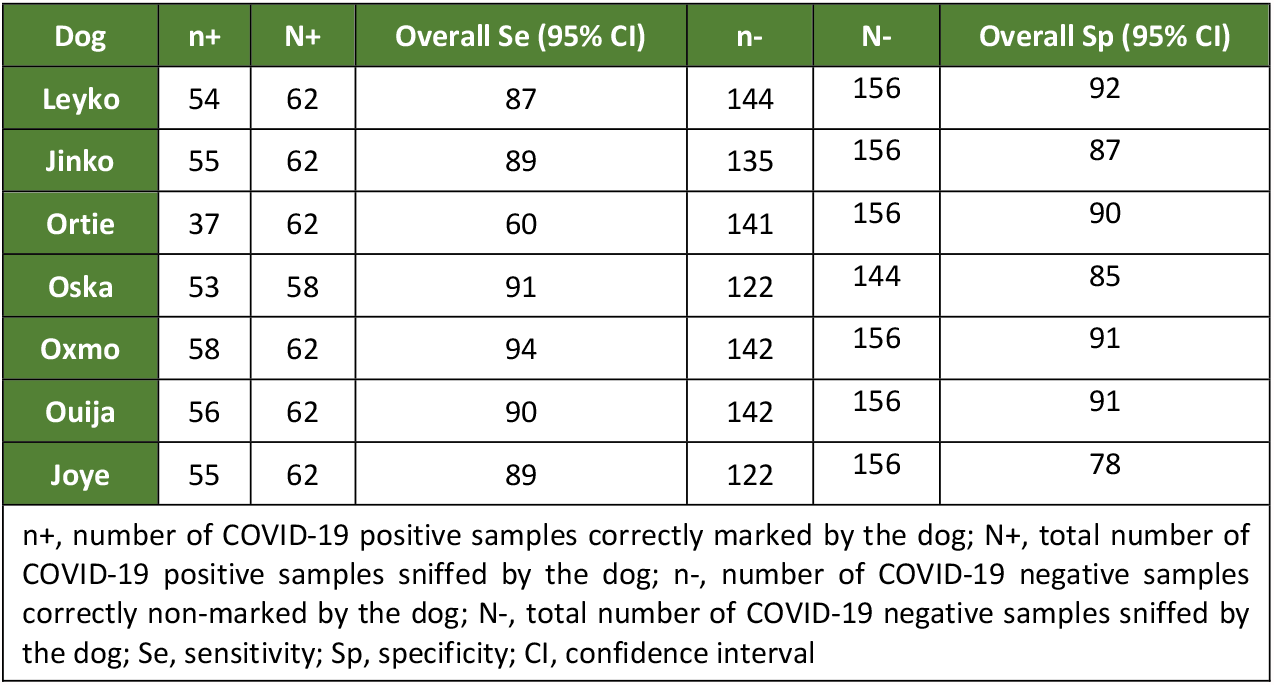
Overall sensitivities and specificities of the 7 dogs calculated from the 218 patients.

| Dog | n+ | N+ | Overall Se (95% CI) | n- | N- | Overall Sp (95% CI) |
| --- | --- | --- | --- | --- | --- | --- |
| Leyko | 54 | 62 | 87 | 144 | 156 | 92 |
| Jinko | 55 | 62 | 89 | 135 | 156 | 87 |
| Ortie | 37 | 62 | 60 | 141 | 156 | 90 |
| Oska | 53 | 58 | 91 | 122 | 144 | 85 |
| Oxmo | 58 | 62 | 94 | 142 | 156 | 91 |
| Ouija | 56 | 62 | 90 | 142 | 156 | 91 |
| Joye | 55 | 62 | 89 | 122 | 156 | 78 |

n+, number of COVID-19 positive samples correctly marked by the dog; N+, total number of COVID-19 positive samples sniffed by the dog; n-, number of COVID-19 negative samples correctly non-marked by the dog; N-, total number of COVID-19 negative samples sniffed by the dog; Se, sensitivity; Sp, specificity; CI, confidence interval

**Table 5.** Sensitivities and specificities for the 7 dogs stratified according to sex of the 218 patients, age, and among patients recruited from AP-HP hospitals (Parisian hospitals) or among patients recruited from GHT Yvelines Nord hospitals.

| Dog* | Females | Males | ≤ 50 years | > 50 years | Foch Hospital | Rambouillet Hospital |
| --- | --- | --- | --- | --- | --- | --- |
| Leyko (87/92) | 84/94 | 90/91 | 86/91 | 90/95 | 100/88 | 90/100 |
| Jinko (89/87) | 87/88 | 90/85 | 86/86 | 90/89 | 84/71 | 91/88 |
| Ortie (60/90) | 65/88 | 55/92 | 59/92 | 62/88 | 74/88 | 60/92 |
| Oska (91/85) | 90/88 | 93/82 | 95/85 | 89/85 | 88/76 | 100/83 |
| Oxmo (94/91) | 90/90 | 97/92 | 95/91 | 92/91 | 95/82 | 90/92 |
| Ouija (90/91) | 87/90 | 94/92 | 86/91 | 92/91 | 100/88 | 80/83 |
| Joye (89/78) | 90/78 | 87/78 | 82/81 | 92/74 | 95/88 | 80/67 |
| *Overall sensitivities/specificities from Table 3 are provided for each dog in parentheses. Numbers are expressed as sensitivities/specificities. |  |  |  |  |  |  |

**Table 6.** Sensitivities and specificities for the 7 dogs among patients free of the most frequently reported health conditions and among patients not using the most frequently reported drugs.

|  | Absence of clinical signs |  |  |  |  | Absence of past or current diseases |  | Not using drugs |  |  |  |  |
| --- | --- | --- | --- | --- | --- | --- | --- | --- | --- | --- | --- | --- |
| Dog* | Dyspnoea | Fatigue | Fever | Dry cough | Muscular pain | Hypertension | Diabetes | Pain killers | Anti-coagulant | Anti-hypertension | Anti-inflammatory | Antibiotics |
| Leyko (87/92) | 93/93 | 89/92 | 89/92 | 86/92 | 93/92 | 85/93 | 85/92 | 90/93 | 88/93 | 85/93 | 86/93 | 85/93 |
| Jinko (89/87) | 90/88 | 88/86 | 89/86 | 90/87 | 91/87 | 93/87 | 85/87 | 94/85 | 90/88 | 89/87 | 88/88 | 85/86 |
| Ortie (60/90) | 52/91 | 61/91 | 66/91 | 58/90 | 63/91 | 68/91 | 57/91 | 62/91 | 62/91 | 62/91 | 56/90 | 56/91 |
| Oska (91/85) | 89/86 | 94/86 | 91/86 | 94/86 | 98/87 | 95/85 | 88/86 | 94/83 | 89/86 | 96/83 | 89/84 | 89/86 |
| Oxmo (94/91) | 93/92 | 91/91 | 94/91 | 94/91 | 95/91 | 95/92 | 91/90 | 96/93 | 95/92 | 96/91 | 92/92 | 92/91 |
| Ouija (90/91) | 90/91 | 85/91 | 91/92 | 94/91 | 88/91 | 90/91 | 87/91 | 92/91 | 93/90 | 91/90 | 92/91 | 86/88 |
| Joye (89/78) | 79/78 | 88/79 | 89/79 | 86/78 | 86/78 | 88/81 | 85/77 | 88/80 | 90/78 | 89/79 | 86/79 | 90/77 |
| *Overall sensitivities/specificities from Table 3 are provided for each dog in parentheses. Numbers are expressed as sensitivities/specificities. |  |  |  |  |  |  |  |  |  |  |  |  |

Table 7 provides PPV and NPV for each dog according to three scenarii according to the risks that an individual is likely to be infected by SARS-CoV-2. If each of the 7 dogs involved in this study does not mark a sample which has 40% of chances or less of coming from a SARS-CoV-2-infected individual, then for all dogs but one (Ortie), the probability that this individual is not SARS-Cov-2-infected (i.e., NPV) is equal to or higher than 91%, and nearly reaches the value of an almost perfect diagnostic tool (97%). Good performances of dogs when not marking a sample are still observed for 3 of the 7 dogs if sniffing a sample which has 50% of chances of coming from a SARS-CoV-2-infected individual (with NPV equal or greater than 90%, close to the 95% reached by an almost perfect diagnostic tool). Performances of dogs when marking a sample are good but to a lesser extent: when the chances of SARS-CoV-2-infection are low (10%), the PPV ranged from 40% to 55% for all dogs (with a PPV of 68% for an almost perfect diagnostic tool). As expected, PPV of dogs when marking a sample increase with increasing probability of SARS-CoV-2-infection, and may be considered as good (ie, ≥ 90%) for 3 out of 7 dogs (Leyko, Oxmo, and Ouija) when the probability of SARS-CoV-2-infection reaches 50%.

**Table 7.** Positive and negative predicted values for the 7 dogs according to the risks that an individual is infected by SARS-Cov2 based on clinical signs and/or prevalence rate in the housing or working area (ranging from 10% to 50% of risks to be infected by SARS-CoV2)

|  | 10% |  | 20% |  | 30% |  | 40% |  | 50% |  |
| --- | --- | --- | --- | --- | --- | --- | --- | --- | --- | --- |
| Dog or almost perfect diagnostic tool* | PPV | NPV | PPV | NPV | PPV | NPV | PPV | NPV | PPV | NPV |
| Almost perfect diagnostic tool (95/95) | 68% | 99% | 83% | 99% | 89% | 98% | 93% | 97% | 95% | 95% |
| Leyko (87/92) | 55% | 98% | 73% | 97% | 82% | 94% | 88% | 91% | 92% | 88% |
| Jinko (89/87) | 43% | 99% | 63% | 97% | 75% | 95% | 82% | 92% | 87% | 89% |
| Ortie (60/90) | 40% | 95% | 60% | 90% | 72% | 84% | 80% | 77% | 86% | 69% |
| Oska (91/85) | 40% | 99% | 60% | 97% | 72% | 96% | 80% | 93% | 86% | 90% |
| Oxmo (94/91) | 54% | 99% | 72% | 98% | 82% | 97% | 87% | 96% | 91% | 94% |
| Ouija (90/91) | 53% | 99% | 71% | 97% | 81% | 96% | 87% | 93% | 91% | 90% |
| Joye (89/78) | 53% | 99% | 50% | 97% | 81% | 96% | 87% | 93% | 80% | 88% |
| *Overall sensitivities/specificities from Table 6 are provided for each dog in parentheses. Numbers are expressed as sensitivities/specificities. |  |  |  |  |  |  |  |  |  |  |

## DISCUSSION

Throughout this validation study set to estimate the individual sensitivity and specificity values of the 7 participating dogs, 218 samples (from 62 COVID-19 positive and 156 COVID-19 negative individuals) from 13 sampling sites were used. The sensitivity values obtained ranged from 87% to 94% for 6 of the dogs, and were above 90% for 3 of them. The dog “Ortie” had a sensitivity of 60% and was therefore not subsequently selected to enter the operational stage on field. Specificity results ranged from 78% to 92%, 6 dogs being above 85% and 4 being above 90%. These numbers appear to be consistent with the ones obtained by Jendrny [4], Eskandari [6], Bjorkman [76], Sarkis [77] and Grandjean [78], who also worked on sweat samples. When using urine samples and then saliva samples, deactivated by detergent, Essler [7] obtained more mixed results, whereas Vesga [5], working on respiratory secretions and saliva, showed sensitivities from 6 dogs ranging between 90% and 98%, and specificities higher than 99%. The World Health Organisation published a document in March 2021 [79] which provides a comprehensive summary of the results obtained in several countries on the subject of canine olfactory detection of COVID-19.

The calculations of sensitivities and specificities in our study used the nasopharyngeal RT-PCR as the “gold standard”, i.e., a detection tool with sensitivity and specificity both equal to 100%. The conclusion of the recently published paper by Axell-House et al. [80], based on a study of over 200 molecular diagnostic tests approved by the Food and Drug Administration (FDA) in the USA for COVID-19, was that many studies trying to present convincing results lack solidity. This is confirmed by Zhang et al., who also add that clinical and public health decisions cannot be solely based on RT-PCR tests [81]. More specifically, Arevalo-Rodriguez et al. quantified a 54% rate of false-positives from RT-PCR tests for COVID-19 in a meta-analysis of nearly 13 000 patients from 34 different studies [82]. The quality of the PCR tests available on the market must be reliable for the training of dogs in olfactory viral detection.

Over time during the cycles, the RT-PCR raises the amount of viral genetic material in the sample of the test to make it detectable. A higher concentration of virus in the sample means a lower number of cycles needed to obtain a detectable amount of genetic material. When performing the RT-PCR, a number of cycles is set (Cycle threshold or Ct) to determine whether a patient is positive or negative. Depending on the material or the laboratory, the range of the scale for the Ct lies between 20 and 45. Up to 30, the rate of false positives by PCR analysis is considered to be very low. Above 30, this rate increases since these performed tests with such high thresholds can detect living material but also genetic fragments, “leftovers” from the infection which do not reflect a SARS-CoV-2 positivity at this time [83]. Similarly, false negatives by PCR analysis are not uncommon (and can reach up to 30% among PCR-negative individuals when tested for the first time [84]). This phenomenon appears to be accentuated as the prevalence of infection decreases in the population [85]. In the USA, the Centre for Disease Control (CDC) recommends using a Ct under 30 to ensure reliable sensitivity values for PCR tests, but manufacturers and laboratories who use them are free to define this number themselves [86].

On top of related public health and ethical issues, such variability in Ct leading therefore to potential high false positive rates became a hindrance to our work on proof of concept during their evolution over the course of the summer 2020 [3]: dogs which did not mark samples identified as being positive, and new criteria to select future samples to train new dogs which must take into account the Ct value of the PCR tests. The dogs initial training (imprinting) and the testing process which determines the individual values for sensitivity and specificity both rely on the accuracy of the current reference test that is the PCR analysis of nasopharyngeal samples.

False positivity on PCR tests may also come from various technical and sampler-dependent issues (such as external contamination during sampling, reagent contamination, cross-reaction with other viruses or remnants of residual genetic material…) [87]. The rates of false positives due to these factors could be over 4% in the United Kingdom [88], and even higher in the USA [89]. Surkova et al. made some suggestions to reduce the risk of false positives: to strengthen quality controls by the laboratory by blindly renewing a small amount of tests in order to rule out any with sub-optimal performance, to take into account pre-test probability, to develop recommendations surrounding the interpretation of results, and to be aware of the clinical and epidemiological meaning of prolonged viral carriage and of the part people in recovery play in the transmission of the disease [87].

In this context, and because elevated false negative and false positive rates strongly complicate the initial training of dogs as it entirely relies on the accuracy of the results from the reference analytical method, we had to narrow the inclusion criteria of positive samples selected for the training stage (pathognomonic clinical symptomatology), and to systematically request for PCR test renewals when the dogs marked a negative sample.

Our study has some strengths in accordance with previous recommendations for detection dog studies [71, 72, 90]. These include different samples being used in the training and validation sessions, randomised sample position, only one sweat sample per individual, samples sniffed only once per dog, double-blinding thanks to a one-way glass, and a large number of recruited individuals (n=218).

In detection dog studies using line-ups, to be confident that dogs detect the studied disease itself and not a characteristic specific to the disease (a confounding factor), disease and disease-free samples in the line-up must be comparable, except for the disease status [36, 72]. COVID-19 negative and positive samples were matched only on hospital. As with observational studies (such as case-control studies), matching COVID-19 positive and negative samples for other potential confounders would have been complicated. This is because the sweat sample had to be sniffed within days after sampling making it impossible to wait for an appropriate matched sample. However, confounding was taken into account in statistical analysis by performing stratification (on sex and age) or restriction (on hospitals, clinical signs, and drugs). Theoretically, if a characteristic was a confounding factor causing high overall sensitivities or specificities, stratification or restriction on one characteristic would have systematically decreased sensitivity and specificity in all the 7 dogs. Although sensitivities slightly decreased after restricting on the absence of diabetes, such overall decrease was not observed for specificities. However, diabetes may exacerbate VOCs related to COVID-19, which would explain the slightly lower sensitivities among individuals free of diabetes. Although the COVID-19 negative and positive samples were matched on hospitals, the two groups were not similar on this characteristic (Table 3). This is because the number of COVID-19 negative samples per COVID-19 positive sample could not have been fixed. After restricting the samples on Foch hospital, the specificity for all the 7 dogs were lower compared to the overall specificity. Because of the low number of COVID-19 negative individuals recruited from Foch hospital, we cannot rule out that these specificities may have lacked some accuracy. However, no such overall decrease in sensitivities were observed among samples collected from Foch hospital. Altogether, the results of our study provide some evidence that the overall sensitivities and specificities were not confounded by confounding bias due to sex, age, hospital, clinical signs, past or current diseases, and used drugs. We however cannot rule out that characteristics other than the ones collected, could possibly cause a residual confounding bias.

The purpose of table 7 was to offer a virtual approach of what the calculated positive and negative predictive values would be for each of the dogs placed for mass detection according to the probability that one individual is infected by SARS-CoV-2 based on clinical signs and/or prevalence rate in the housing or working area (ranging from 10% to 50%). These positive and negative predictive values for each dog were also compared to the values obtained from an “almost perfect” test (95% sensitivity and specificity), whichever it may be. Table 7 shows that, while on one hand the positive predictive value of the dog is lower than the “almost perfect” test (although it does rise when the prevalence rate rises), the negative predictive value on the other hand is similar to the reference.

This study aimed to confirm the hypothesis we previously suggested [3] that olfactory detection dogs trained to detect the presence of SARS-CoV-2 can be a promising alternative detection tool in settings where conventional diagnostic tools are not available or difficult to implement. This study is also an essential and necessary step before conducting the following subsequent investigations. A first one would be to assess the COVID-19 detection dog’s behaviour in the presence of other diseases, such as viral respiratory diseases or comorbidities such as diabetes. A second one would be to assess whether the trained dogs can detect SARS-CoV-2 infection before a sample is positive by using the PCR method (hypothesis supported by Essler et al. [7]). A third one would to assess the effect of the simplification of the smelling process by the dog (for instance, by reducing the duration of contact with the swabs for axillary samples or the use of olfactory test line-ups faster to set-up and to clean) on dogs’ performance. A fifth one would be to assess the dog’s performance in a population of asymptomatic or paucisymptomatic COVID-19 patients, who represent the population at risk of dissemination in a mass detection framework.

It is now necessary to standardize an optimal training method for large numbers of scent dogs for COVID-19, and infrastructural supporting COVID-19 scent dogs deployments. This has led the World Health Organization to recently set up international working groups for the different areas concerned.

## CONCLUSION

This study, along with other studies in COVID-19 canine detection, supports the hypothesis of strong canine olfactory abilities to detect infection with SARS-CoV-2 on individuals as well as the one of the presence of a specific Volatil Organic Compounds profile associated with the portage of the virus. It also provides priceless information on how to improve training methods and optimize the potential use of such dogs in future mass-detection campaigns. The use of dog for COVID-19 detection have the following advantages over conventional approaches to detect the presence of SARS-CoV-2: non-invasive and cheap method with a result immediately obtained.

## Data Availability

All the data described in this article are available on request

